# Generalized anxiety disorder, depressive symptoms and sleep quality during COVID-19 epidemic in China: a web-based cross-sectional survey

**DOI:** 10.1101/2020.02.19.20025395

**Authors:** Yeen Huang, Ning Zhao

## Abstract

**Background:** China has been severely affected by COVID-19 (Corona Virus Disease 2019) since December, 2019. This study aimed to assess the population mental health burden during the epidemic, and to explore the potential influence factors.

**Methods:** Using a web-based cross-sectional survey, we collected data from 7,236 self-selected volunteers assessed with demographic information, COVID-19 related knowledge, Generalized Anxiety Disorder-7 (GAD-7), Center for Epidemiology Scale for Depression (CES-D), and Pittsburgh Sleep Quality Index (PSQI). Logistic regressions were used to identify influence factors associated with mental health problem.

**Results:** Of the total sample analyzed, the overall prevalence of GAD, depressive symptoms, and sleep quality were 35.1%, 20.1%, and 18.2%, respectively. Young people reported a higher prevalence of GAD and depressive symptoms than older people (*P*<0.001). Compared with other occupational group, healthcare workers have the highest rate of poor sleep quality (*P*<0.001). Multivariate logistic regression showed that age (< 35 years) and times to focus on the COVID-19 (≥ 3 hours per day) were associated with GAD, and healthcare workers were associated with poor sleep quality.

**Conclusions:** Our study identified a major mental health burden of the public during COVID-19 epidemic in China. Young people, people who spent too much time on the epidemic, and healthcare workers were at high risk for mental illness. Continuous surveillance and monitoring of the psychological consequences for outbreaks should become routine as part of preparedness efforts worldwide.

## 1. Introduction

COVID-19 (Corona Virus Disease 2019, also known as 2019-nCoV), a cluster of acute respiratory illness with unknown causes, occurred in Wuhan, Hubei Province, China since December 2019 (Wuhan Municipal Health Commission, 2020; Paules et al., 2020; Wang et al., 2020). As of February 15, 2020, a total of 66,580 COVID-19 cases in China have been confirmed and 1,524 Chinese died from the disease. Internationally, sporadic cases exported from Wuhan were reported in 25 countries (such as 334 cases in Japan, and 67 cases in Singapore), 5 continents, and 1 international conveyance (218 cases in “Diamond Princess”) (World Health Organization, 2020a). On January 23, Wuhan city closed all access routes to stop the spread of disease. Seven days later, the World Health Organization (WHO) declared the COVID-19 as a Public Health Emergency of International Concern (PHEIC) (World Health Organization, 2020b).

In addition to causing physical damage, COVID-19 also has a serious impact on the mental health of the public. In January 20, China confirmed human-to-human transmission of COVID-19 and some medical staff in Wuhan have been infected (XINHUANET, 2020). Since then, the public has shown anxiety-related behaviors, causing a significant shortage of medical masks and alcohol across the country. On the night of January 31, due to a news release that “Shuanghuanglian” oral liquid could suppress COVID-19 (People’s daily of China, 2020), the public rushed to pharmacies overnight to buy this drug. In addition, many front-line medical staff work more than 16 hours a day on average, causing them to not get enough sleep. Unfortunately, a 37-year-old Japanese government worker who was in charge of isolated returnees from Wuhan was found to have died from apparent suicide (The Japan Times, 2020).

Evidence indicated that COVID-19 is a distinct clade from the betacoronaviruses related to human severe acute respiratory syndrome (SARS) and Middle East respiratory syndrome (MERS) (Zhu et al., 2020). Several studies showed that mental health problems could occur in both healthcare workers and SARS survivors during the SARS epidemic (Lee et al., 2007; Lu et al., 2006; McAlonan et al., 2007). Post-traumatic stress disorder (PTSD) and depressive disorders were the most prevalent long-term psychological condition (Mak et al., 2009). Similar results have been reported in the previous study of MERS (Lee et al., 2018). Based on the above research evidence, we have reason to speculate that the psychological condition of the public may also be affected during COVID-19 epidemic.

Therefore, using a web-based cross-sectional study, we aimed to assess the mental health burden of Chinese population during COVID-19 epidemic, and to explore the potential influence factors. We hope our study findings will provide data support for the targeted interventions on psychological health in Chinese population during the epidemic.

## 2. Methods

### 2.1 Study design and participants

To prevent the spread of SARS-CoV-2 (Severe Acute Respiratory Syndrome Coronavirus 2) through droplets or contact, we used a web-based cross-sectional survey based on the National Internet Survey on Emotional and Mental Health (NISEMH) to collected data, NISEMH is an ongoing, online health-related behavior survey of Chinese population. This web-based survey of the COVID-19 was broadcasted on the Internet through the WeChat public platform and the mainstream media. All Chinese people using WeChat or other social tools may see this survey, and answered the questionnaire by scanning the two-dimensional barcodes of the questionnaire address or clicking the relevant link. To encourage the recruitment of potential participants, all participants in the survey would receive a report on their mental health after completing the evaluation. This web-based questionnaire was completely voluntary and non-commercial.

### 2.2 Data collection

Participants answered the questionnaires anonymously on the Internet from February 3, 2020 to February 17, 2020. All subjects reported their demographic data, COVID-19 related information, and three standardized questionnaires, which assessed their generalized anxiety disorder (GAD), depressive symptoms, and sleep quality. In order to ensure the quality of survey, we have set the response range of some items (e.g., the age range was limited to 6-80 years old, some items needed to be answered in reverse) and encouraged participants to answer carefully through questionnaire explanations. In addition, questionnaires that were completed <1 minute or >60 minutes would be excluded from analysis. Finally, a total of 7,236 participants who completed the questionnaires (response rate of 85.3%) were included in the analysis.

### 2.3 Ethical statement

This study was conducted in accordance with the Declaration of Helsinki, and was approved by the Ethics Committee of Huazhong University of Science and Technology Union Shenzhen Hospital. Electronic informed consent was obtained from each participants prior to starting the investigation. Participant could withdraw from the survey at any moment without providing any justification.

### 2.4 Measures

#### 2.4.1 Demographic information

Demographic variables included gender (male or female), age, and occupation. Occupation included the following four types: (1) Healthcare workers, which included doctors, nurses, and health-related administrators; (2) Enterprise or institution workers, which consisted of enterprise employees, national/provincial/municipal institution workers, and other relevant staff; (3) Teachers or students, which included teachers or students from universities, middle schools, or elementary schools; and (4) Others, which consisted of freelancers, retiree, social worker, and other relevant staff.

#### 2.4.2 COVID-19 related knowledge

This section was evaluated by two items: (1) Times to focus on the COVID-19, which measured the average time spent focusing on the COVID-19 epidemic information every day; (2) Knowledge of the COVID-19, which assessed based on the following six judgment questions about COVID-19 related knowledge: a. Inhalation of droplets from sneezing, coughing, or talking of an infected person could cause infection; b. Contact with something contaminated by an infected person could lead to infection; c. The incubation period of the virus does not exceed 14 days; d. Contact with an asymptomatic person might also lead to infection; e. There are already targeted drugs that could cure the disease; f. Taking “Shuanghuanglian Oral Liquid” could prevent infection of this disease. Of the above six questions, one point was given for correct answers, and no points were given for incorrect or uncertain answers. Participants with scores ≥5 points, equal to 4 points, and ≤3 points were considered to be quite understand, generally understand, and do not understand.

#### 2.4.3 Generalized anxiety disorder

We used Chinese version of GAD-7 (Generalized Anxiety Disorder-7) scale to assess subject’s anxiety symptoms. The GAD-7 has been previously used in Chinese populations, and found to have good reliability (Cronbach’s alpha=0.90) (Tong et al., 2016; Wang et al., 2018). Seven items assess the frequency of anxiety symptoms over the past two weeks on a 4-point Liker-scale ranging from 0 (never) to 3 (nearly every day). The total score of GAD-7 ranged from 0 to 21, with increasing scores indicated a more severe functional impairments as a result of anxiety (Spitzer et al., 2006). For the purpose of this study, we defined a GAD-total score of 9 points or greater as the presence of anxiety symptoms (Wang et al., 2018).

#### 2.4.4 Depressive symptoms

The Center for Epidemiology Scale for Depression (CES-D) in Chinese version was used to identify whether participants had depressive symptoms (Zhang et al., 2010), and the Chinese version of this scale has been validated and extensively utilized in Chinese population (Zhang et al., 2010; Zhang and Li, 2011). Twenty items assess the frequency of depressive symptoms over the past two weeks on a 4-point Liker-scale ranging from 0 (rarely or none of the time) to 3 (most or all of the time). The score range of the CES-D is 0-60 points, and higher scores indicated more severe depressive symptomatology (Radloff, 1977). In our study, CES-D scores greater than 28 points indicated depressive symptoms.

#### 2.4.5 Sleep quality

The Chinese version of the PSQI (Pittsburgh Sleep Quality Index) scale was used to assess the subject’s sleep quality over the past two weeks (Liu et al., 1996). The PSQI scale contains seven components (subjective sleep quality, sleep duration, sleep latency, habitual sleep efficiency, use of sleep medications, sleep disturbance, and daytime dysfunction), and the score for each component ranging from 0 to 3 points. The global PSQI score ranges from 0 to 21, with higher scores indicated more severe sleep disorder (Buysse et al., 1989). The Chinese version of PSQI has been demonstrated to be reliable and valid in Chinese population (Liu et al., 1996), a global PSQI score greater than 7 points indicated poor sleep quality.

#### 2.4.6 Statistical analysis

First, descriptive analyses were conducted to describe the demographic characteristics and COVID-19 related knowledge in Chinese population. Second, the prevalence of GAD, depressive symptoms, and sleep quality stratified by gender, age,and occupation were reported, and Chi-square test (χ^*2*^) was used to compare the differences between groups. Third, univariate and multivariate logistic regression models were performed to explore potential influence factors for GAD, depressive symptoms, and sleep quality during COVID-19 epidemic. Odds ratio (*OR*), adjusted odds ratio (*AOR*), and 95% confidence interval (95% *CI*) were obtained from logistic regression models. All data were analyzed using Statistical Package for Social Sciences (SPSS) version 24.0. *P*-values of less than 0.05 were considered statistically significant (2-sided tests).

## 3. Results

### 3.1 Demographic characteristics

The characteristics of participants were shown in Table 1. Of the 7,236 sample analyzed, 3,284 (45.4%) were males and 3,952 (54.6%) were females, and the mean (standard deviation) age of the participants was 35.3±5.6 years. Among these samples, 2,250 (31.1%) of participants were healthcare workers, 3,155 (43.6%) participants focused on the COVID-19 for 3 hours or more every day, and 5,702 (78.8%) participants were quite understand for the COVID-19.

**Table 1.** Demographic characteristics of study sample (N=7,236).

| Variable | n (%) |
| --- | --- |
| **Total** | 7236 (100.0) |
| **Gender** |  |
| Male | 3284 (45.4) |
| Female | 3952 (54.6) |
| **Age (Mean±SD)** | 35.3±5.6 |
| < 35 years | 3155 (43.6) |
| ≥ 35 years | 4081 (56.4) |
| **Occupation** |  |
| Healthcare workers<sup>a</sup> | 2250 (31.1) |
| Enterprise or institution workers<sup>b</sup> | 1809 (25.0) |
| Teachers or students<sup>c</sup> | 1404 (19.4) |
| Others<sup>d</sup> | 1773 (24.5) |
| **Times to focus on the COVID-19<sup>e</sup>** |  |
| <1 hour | 1454 (20.1) |
| 1-2 hours | 2627 (36.3) |
| ≥3 hours | 3155 (43.6) |
| **Knowledge of the COVID-19** |  |
| Do not understand (score ≤3 points) | 398 (5.5) |
| General understand (score 4 points) | 1136 (15.7) |
| Quite understand (score ≥5 points) | 5702 (78.8) |

Abbreviations: n, number; SD, Standard deviation; COVID-19, 2019 Corona Virus Disease.
<sup>a</sup>Included doctors, nurses and health administrators.
<sup>b</sup>Included enterprise employees, national/provincial/municipal institution workers and other relevant staff.
<sup>c</sup>Included teachers or students from universities, middle schools, or elementary schools.
<sup>d</sup>Includer freelancers, retiree, social worker and other relevant staff.
<sup>e</sup>Average time spent focusing on the COVID-19 epidemic information every day.

### 3.2 Prevalence of GAD, depressive symptoms, and sleep quality during COVID-19 epidemic stratified by gender, age, and occupation

The prevalence of GAD, depressive symptoms, and sleep quality stratified by gender, age, and occupation were shown in Table 2, Table 3, and Table 4, respectively. The overall prevalence of GAD, depressive symptoms, and sleep quality were 35.1%, 20.1%, and 18.2%, respectively. There was no statistically significant difference in the prevalence of GAD, depressive symptoms, and sleep quality by gender (*P*>0.05), as shown in Table 2. As shown in Table 3, the prevalence of GAD and depressive symptoms was significantly higher in participants younger than 35 years than in participants aged 35 years or older (*P*<0.001). Compared with other occupational groups, healthcare workers (23.6%) reported the highest rate of poor sleep quality (*P*<0.001), as shown in Table 4.

**Table 2.** Prevalence of GAD, depressive symptoms, and sleep quality during COVID-19 epidemic in Chinese population stratified by gender (N=7,236).

| Variables | Total (N=7236)<br>n (%) | Male (N=3284)<br>n (%) | Female (N=3952)<br>n (%) | $\chi^2$ | P-value |
| --- | --- | --- | --- | --- | --- |
| **GAD<sup>a</sup>** |  |  |  | 2.89 | 0.089 |
| No | 4696 (64.9) | 2092 (63.7) | 2394 (65.9) |  |  |
| Yes | 2540 (35.1) | 1192 (36.3) | 1348 (34.1) |  |  |
| **Depressive symptoms<sup>b</sup>** |  |  |  | 3.67 | 0.055 |
| No | 5782 (79.9) | 2625 (80.0) | 3155 (79.8) |  |  |
| Yes | 1454 (20.1) | 657 (20.0) | 797 (20.2) |  |  |
| **Sleep quality<sup>c</sup>** |  |  |  | 2.59 | 0.108 |
| Good | 5919 (81.8) | 2660 (81.0) | 3259 (82.5) |  |  |
| Poor | 1317 (18.2) | 624 (19.0) | 693 (17.5) |  |  |

Abbreviations: n, number, GAD, generalized anxiety disorder. <sup>a</sup>GAD was defined as individuals who scored $\geq 9$ points. <sup>b</sup>Depressive symptoms included individuals who scored $\geq 28$ points. <sup>c</sup>Poor sleep quality was defined as individuals who scored $> 7$ points.

**Table 3.** Prevalence of GAD, depressive symptoms, and sleep quality during COVID-19 epidemic in Chinese population stratified by age (N=7,236).

| Variables | Total<br>(N=7236)<br>n (%) | Age < 35 year<br>(N=3155)<br>n (%) | Age ≥ 35 year<br>(N=4081)<br>n (%) | $\chi^2$ | P-value |
| --- | --- | --- | --- | --- | --- |
| **GAD<sup>a</sup>** |  |  |  | 20.67 | <0.001 |
| No | 4696 (64.9) | 1956 (62.0) | 2740 (67.1) |  |  |
| Yes | 2540 (35.1) | 1199 (38.0) | 1341 (32.9) |  |  |
| **Depressive symptoms<sup>b</sup>** |  |  |  | 13.91 | <0.001 |
| No | 5782 (79.9) | 2458 (77.9) | 3324 (81.5) |  |  |
| Yes | 1454 (20.1) | 697 (22.1) | 757 (18.5) |  |  |
| **Sleep quality<sup>c</sup>** |  |  |  | 0.58 | 0.446 |
| Good | 5919 (81.8) | 2575 (81.6) | 3344 (81.9) |  |  |
| Poor | 1317 (18.2) | 580 (18.4) | 737 (18.1) |  |  |

Abbreviations: n, number, GAD, generalized anxiety disorder. <sup>a</sup>GAD was defined as individuals who scored ≥ 9 points. <sup>b</sup>Depressive symptoms included individuals who scored ≥ 28 points. <sup>c</sup>Poor sleep quality was defined as individuals who scored > 7 points.

**Table 4.** Prevalence of GAD, depressive symptoms, and sleep quality during COVID-19 epidemic in Chinese population stratified by Occupation (N=7,236).

| Variables | Total<br>(N=7236)<br>n (%) | Healthcare<br>workers<br>(N=2250)<br>n (%) | Enterprise or<br>institution<br>workers (N=1809)<br>n (%) | Teachers or<br>students<br>(N=1404)<br>n (%) | Others<br>(N=1773)<br>n (%) | $\chi^2$ | P-value |
| --- | --- | --- | --- | --- | --- | --- | --- |
| **GAD<sup>a</sup>** |  |  |  |  |  | 2.36 | 0.501 |
| No | 4696 (64.9) | 1448 (64.4) | 1179 (65.2) | 911 (64.9) | 1158 (65.3) |  |  |
| Yes | 2540 (35.1) | 802 (35.6) | 630 (34.8) | 493 (35.1) | 615 (34.7) |  |  |
| **Depressive symptoms<sup>b</sup>** |  |  |  |  |  | 2.71 | 0.439 |
| No | 5782 (79.9) | 1804 (80.2) | 1445 (79.9) | 1109 (79.0) | 1424 (80.3) |  |  |
| Yes | 1454 (20.1) | 446 (19.8) | 364 (20.1) | 295 (21.0) | 349 (19.7) |  |  |
| **Sleep quality<sup>c</sup>** |  |  |  |  |  | 98.82 | <0.001 |
| Good | 5919 (81.8) | 1719 (76.4) | 1579 (87.3) | 1203 (85.7) | 1418 (80.5) |  |  |
| Poor | 1317 (18.2) | 531 (23.6) | 230 (12.7) | 201 (14.3) | 355 (20.0) |  |  |

Abbreviations: n, number, GAD, generalized anxiety disorder.
<sup>a</sup>GAD was defined as individuals who scored $\geq 9$ points.
<sup>b</sup>Depressive symptoms included individuals who scored $\geq 28$ points.
<sup>c</sup>Poor sleep quality was defined as individuals who scored $> 7$ points.

### 3.3 Association of influence factors with GAD, depressive symptoms, and sleep quality during COVID-19 epidemic

The associations of potential influence factors with GAD, depressive symptoms, and sleep quality during COVID-19 epidemic were presented in Table 5. In the univariate logistic regression models, age (*OR*=1.77, 95% *CI*: 1.38-1.95) and times to focus on the COVID-19 (*OR*=1.91, 95% *CI*: 1.77-2.15) were significantly associated with GAD in Chinese population. Similarly, age were associated with depressive symptoms (*OR*=1.80, 95% *CI*: 1.35-2.01), but not with sleep quality (*OR*=0.69, 95% *CI*: 0.35-1.05). Occupation was related to sleep quality during COVID-19 epidemic in Chinese population (*OR*=1.48, 95% *CI*: 1.15-1.95).

**Table 5.** Results of univariate and multivariate logistic regression analyses (N=7,236).

| Variables | GAD |  | Depressive symptoms |  | Sleep quality |  |
| --- | --- | --- | --- | --- | --- | --- |
|  | *OR (95% CI)* | *AOR (95% CI)* | *OR (95% CI)* | *AOR (95% CI)* | *OR (95% CI)* | *AOR (95% CI)* |
| **Gender** |  |  |  |  |  |  |
| Male | 1.00 | 1.00 | 1.00 | 1.00 | 1.00 | 1.00 |
| Female | 1.32 (0.90-1.69) | 1.22 (0.86-1.64) | 1.30 (0.82-2.07) | 1.24 (0.77-1.99) | 0.89 (0.57-1.39) | 0.82 (0.52-1.29) |
| **Age** |  |  |  |  |  |  |
| < 35 years | **1.77 (1.38-1.95)\*** | **1.65 (1.49-2.02)\*** | **1.80 (1.35-2.01)\*** | **1.77 (1.58-2.07)\*** | 0.69 (0.35-1.05) | 0.68 (0.42-1.11) |
| ≥ 35 years | 1.00 | 1.00 | 1.00 | 1.00 | 1.00 | 1.00 |
| **Occupation** |  |  |  |  |  |  |
| Healthcare workers<sup>a</sup> | 1.30 (0.83-2.04) | 1.30 (0.82-2.08) | 1.15 (0.67-1.99) | 1.02 (0.58-1.81) | **1.48 (1.15-1.95)\*** | **1.32 (1.18-1.88)\*** |
| Enterprise or institution workers<sup>b</sup> | 0.85 (0.52-1.38) | 0.91 (0.55-1.49) | 0.80 (0.44-1.47) | 0.80 (0.44-1.49) | 0.60 (0.33-1.11) | 0.59 (0.32-1.10) |
| Teachers or students<sup>c</sup> | 1.51 (0.91-2.53) | 1.41 (0.80-2.50) | 1.24 (0.67-2.31) | 0.94 (0.47-1.88) | 0.69 (0.35-1.35) | 0.87 (0.42-1.82) |
| Others<sup>d</sup> | 1.00 | 1.00 | 1.00 | 1.00 | 1.00 | 1.00 |
| **Times to focus on the COVID-19<sup>e</sup>** |  |  |  |  |  |  |
| <1 hour | 1.00 | 1.00 | 1.00 | 1.00 | 1.00 | 1.00 |
| 1-2 hours | 0.96 (0.59-1.57) | 1.01 (0.61-1.64) | 0.71 (0.40-1.27) | 0.74 (0.41-1.32) | 0.90 (0.50-1.62) | 0.81 (0.44-1.49) |
| ≥3 hours | **1.91 (1.77-2.15)\*** | **1.83 (1.53-2.19)\*** | 0.98 (0.57-1.68) | 1.11 (0.63-1.93) | 1.18 (0.68-2.07) | 1.02 (0.57-1.82) |
| **Knowledge of the COVID-19** |  |  |  |  |  |  |
| Do not understand | 1.00 | 1.00 | 1.00 | 1.00 | 1.00 | 1.00 |
| General understand | 0.73 (0.32-1.71) | 0.68 (0.29-1.60) | 0.97 (0.32-2.97) | 0.90 (0.29-2.76) | 1.06 (0.35-3.21) | 0.92 (0.30-2.82) |
| Quite understand | 0.93 (0.45-1.93) | 0.80 (0.38-1.69) | 1.30 (0.49-3.47) | 1.12 (0.42-3.02) | 1.29 (0.48-3.42) | 1.15 (0.42-3.14) |

Abbreviations: GAD, generalized anxiety disorder; *OR*, odds ratio; *AOR*, adjusted odds ratio; 95% *CI*, 95% confidence interval; COVID-19, 2019 Corona Virus Disease. <sup>a</sup>Included doctors, nurses and health administrators. <sup>b</sup>Included enterprise employees, national/provincial/municipal institution workers and other relevant staff. <sup>c</sup>Included teachers or students from universities, middle schools, or elementary schools. <sup>d</sup>Includer freelancers, retiree, social worker and other relevant staff. <sup>e</sup>Average time spent focusing on the COVID-19 epidemic information every day.
\* $P<0.001$ .

In the multivariate logistic regression models, the above associations have weakened but there were still statistical difference. Participants under 35 years were more likely to have GAD than those 35 years and older (*AOR*=1.65, 95% *CI*: 1.49-2.02). Besides, participants who were concerned about the COVID-19 epidemic for 3 hours or more were more likely to develop GAD than those less than 1 or 2 hours (*AOR*=1.83, 95% *CI*: 1.53-2.19). Similarly, participants under 35 were associated with higher risk for depressive symptoms than those 35 years and older (*AOR*=1.77, 95% *CI*: 1.58-2.07). Compared with other occupation participants, healthcare workers were more likely to report poor sleep quality (*AOR*=1.32, 95% *CI*: 1.18-1.88).

## 4. Discussion

Our web-based study show a high prevalence of GAD and poor sleep quality in the Chinese population during COVID-19 epidemic. Anxiety symptoms were more likely to occur in people younger than 35 years and those who spent too much time focusing on the epidemic. Compared with other professions, healthcare workers were associated with higher risk for poor sleep quality. Our findings provided data support for accurately understand the source of public’s panic during COVID-19 epidemic.

The data in this study suggested public’s levels of anxiety-related symptoms increase when a major infectious diseases occurred. Similar to the psychological burden caused by SARS (Su et al., 2007), we found that one in three participants showed anxiety disorders, and this mood was not different between male and female during COVID-19 epidemic, which was different from previous research that women were more likely to have anxiety than men (Guo et al., 2016; Gao et al., 2020). In addition, nearly one in five participants had depressive symptoms and sleep problems, indicating that the uncertainty of the epidemic progression would cause greater psychological pressure on the public. The possible reason for these mental problems may be related to the “hypochondriac concerns” (worry about being infected) (Furer et al., 1997) and feared that the epidemic was hard to control.

After multivariate logistic regression analyses, we found that age and times to focus on COVID-19 may be potential risk factors for the psychological problems of the public. Younger participants (< 35 years) were more likely to develop anxiety and depressive symptoms during COVID-19 epidemic than older participants (≥ 35 years). Our results were similar to those of a previous study in Taiwan during SARS outbreak (Su et al., 2007). In addition, we assessed the average time participants spent focusing on the COVID-19 epidemic each day, and found that people who spent too much time on the epidemic (≥ 3 hours) were more likely to develop anxiety symptoms. The manifestation of this panic mood may be related to the body’s normal protective response to the stress caused by the epidemic (Maunder et al., 2003).

Since January 20, 2020, Zhong Nanshan (the renowned Chinese respiratory expert who discovered the SARS virus) confirmed that there must be human-to-human transmission of COVID-19 (XINHUANET, 2020), more than 20,000 medical staff gave up the Spring Festival holiday and voluntarily applied to support the epidemic in Hubei Province (National Health Commission of the People’s Republic of China, 2020). Meanwhile, most healthcare workers in China have returned to work to cope with the further development of the disease. Our findings showed that nearly one in four healthcare workers have sleep problems, which was significantly higher than other occupational group. One possible reason is that the working intensity and time of healthcare workers will increase in the face of severe epidemic (such as SARS and MERS), resulting in them not having enough time to rest, and prone to chronic stress and psychological distress (Lu et al., 2006; Lee et al., 2018; McAlonan et al., 2007). In severe cases, a post-traumatic stress disorder (PTSD) symptoms may even occur, which is highly correlated with poor sleep (Kobayashi et al., 2007).

Fortunately, the Chinese government has taken many strong national measures in time to avoid further spread of the COVID-19 epidemic, including requiring uninfected people to isolate themselves at home, prohibiting all gathering activities, and forcing everyone to wear medical masks to enter public places. However, there is still lack of relevant research on the targeted intervention of the public’s psychological problems during the COVID-19 epidemic. We filled this research gap by analyzing the prevalence of psychological issue in Chinese population stratified by demographic characteristics and exploring related influential factors. Several appropriate interventions are recommended as follows: First, particular effect should be direct to vulnerable populations which include the suspected and diagnosed patients, young people, and healthcare workers, especially physicians and nurses working directly with patients or quarantined people. Second, try to control the time of receiving relevant information no more than two hour every day, focus only on the necessary information (such as facts and data) and avoid receiving too many harmful rumors (Grein et al., 2000). Third, maintain normal work and rest as much as possible, exercise regularly to promote sleep quality, and do not pay too much attention to epidemic information before going to sleep.

This study has several limitations. First, the data and relevant analyses presented here were derived from a cross-sectional design, it is difficult to make causal inferences. Second, the study was limited to COVID-19 epidemic, we used web-based survey method to avoid possible infections. However, this leading to the sampling of our study was voluntary and conducted by online system. Therefore, the possibility of selection bias should be considered. Third, due to the sudden occurrence of the disaster, we were unable to assess an individual’s psychiatric conditions before the outbreak.

## 5. Conclusion

In conclusion, we identified a major mental health burden of the public during COVID-19 epidemic in China, and young people, people who spent too much time on the epidemic, and healthcare workers were at a high risk of displaying psychological issues. Previously, when SARS occurred in China, the awareness regarding public’s mental health related to the epidemic was low, and no targeted psychological guidelines available to the public in need during the pandemic period. Therefore, ongoing surveillance and monitoring of the psychological consequences for outbreaks of epidemic-potential, life-threatening diseases, establishing early targeted mental health interventions, should become routine as part of preparedness efforts worldwide.

## Data Availability

Data available if required

## Author statement contributors

Ning Zhao conceptualized and designed the study, review and revised the manuscript, and approved the final manuscript as submitted. Yeen Huang designed the data collection instruments, coordinated and supervised data collection, carried out the initial analyses, and interpreted the data, drafted the initial manuscript, and approved the final manuscript as submitted. Ning Zhao and Yeen Huang agree to be accountable for all aspects of the study.

## Acknowledgements

The authors would like to thank all the participants in our study. In addition, we express our heartfelt respect to all healthcare workers who are fighting the epidemic on the front line. Finally, we thank Ms. Qiaohong Chen for providing professional language help.

